# Incidence and survival of pediatric and adult hepatocellular carcinoma, United States, 2001-2020

**DOI:** 10.1101/2024.03.25.24304564

**Authors:** Azlann Arnett, David A. Siegel, Shifan Dai, Trevor D. Thompson, Jennifer Foster, Erika J. di Pierro, Behnoosh Momin, Philip J. Lupo, Andras Heczey

**Author notes:** **Corresponding Author:** Andras Heczey MD; Address: 1102 Bates Avenue C1760.10, Houston, TX, 77025. **Disclaimer:** The findings and conclusions in this report are those of the authors and do not necessarily represent the official position of the Centers for Disease Control and Prevention.

## Abstract

**Importance:** Hepatocellular carcinoma accounts for approximately 80% of liver neoplasms. Globally, hepatocellular carcinoma ranks as the third most lethal cancer, with the number of deaths expected to further increase by 2040. In adults, disparities in incidence and survival are well described while pediatric epidemiology is not well characterized.

**Objective:** To describe incidence and survival for pediatric (ages 0-19 years) hepatocellular carcinoma cases and compare these measures to adults (ages ≥20 years) diagnosed with hepatocellular carcinoma. We evaluated demographic factors and clinical characteristics that influence incidence and outcomes.

**Design:** Population-based cohort study.

**Setting:** Incidence data from the US Cancer Statistics database from 2003 to 2020 and 5-year relative survival from the National Program of Cancer Registries from 2001 to 2019, covering 97% and 83% of the US population, respectively.

**Participants:** 355,349 US Cancer Statistics and 257,406 the National Program of Cancer Registries patients were identified using ICD-O-3 C22.0 and 8170-5 codes.

**Main Outcomes and Measures:** Incidence annual percent change (APC) and average APC (AAPC) using joinpoint regression. Five-year relative survival. All-cause survival estimated using multivariate Cox modeling. Corresponding 95% confidence intervals (CI) were calculated.

**Results:** Incidence rate per 100,000 persons was 0.056 (95%CI:0.052-0.060) for pediatric cases and 7.793 (7.767-7.819) for adults. Incidence was stable in the pediatric population (0.3 AAPC, −1.1-1.7). In contrast, after periods of increase, incidence declined in adults after 2015 (−1.5 APC). Relative survival increased over time for both pediatric and adult ages and was higher for children and adolescents (46.4%, 95%CI:42.4-50.3) than adults (20.7%, 95%CI:20.5-20.9) overall and when stratified by stage. Regression modeling showed that non-Hispanic Black race and ethnicity was associated with higher risk of death in children and adolescents (1.48, 95%CI:1.07-2.05) and adults (1.11, 95%CI:1.09-1.12) compared to non-Hispanic white race and ethnicity.

**Conclusions and Relevance:** Between 2003 and 2020 in the United States, pediatric incidence was stable while incidence in adults began to decline after 2015. Survival was higher across all stages for children and adolescents compared to adults. Non-Hispanic Black race and ethnicity showed a higher risk of death for both age groups. Further studies could explore the factors that influence these outcome disparities.

**KEY POINTS:** *Question:* How does incidence and survival compare between pediatric and adult hepatocellular carcinoma (HCC)?

*Findings:* In contrast to adults, pediatric incidence rates of HCC were stable, and no demographic risk factors for incidence were identified. Pediatric HCC survival was higher than adults. Non-Hispanic Black race and ethnicity, small metropolitan county, and non-fibrolamellar histology, were risk factors in both pediatric and adult groups.

*Meaning:* Non-Hispanic Black patients and those living in smaller metropolitan and non-metropolitan areas irrespective of age might benefit from further research in outcome disparities, provider education, and prospective studies to maximize outcomes through effective risk-adapted management strategies.

## INTRODUCTION

Hepatocellular carcinoma (HCC) is the most common type of liver cancer worldwide. HCC has the sixth highest incidence and ranks third for cancer-related deaths with 1.4 million new cases and 1.3 million deaths per year predicted by 2040^1^. In children, HCC typically occurs in the absence of cirrhosis and is frequently associated with predisposing conditions while in adults, hepatitis C virus (HCV), alcoholic liver disease, and metabolic dysfunction-associated steatosis liver disease (MASLD) drive disease pathogenesis^2–5^. Overall, pediatric and adult HCC have comparable histology and are classified similarly though potential biological differences remain to be fully understood^6,7^.

HCC epidemiology has been well characterized in adults but is not well described in children. Reports from the Surveillance Epidemiology and End Results (SEER) database, which covered <30% of the US population, the incidence of pediatric HCC is 0.5-0.59 per million persons^2–4^. Compared to adult studies, analyses of racial and ethnic disparities in pediatric HCC have reported inconsistent results^2–5,8^. Thus, there is a need for in-depth studies with high population coverage to contextualize results from case series and other small sample studies.

We analyze pediatric and adult HCC incidence and outcome data sourced from the United States Cancer Statistics (USCS) and National Program Cancer Registries (NPCR) databases, which cover 97% and 83% of the population, respectively. We compare results by age to highlight differences and similarities that may inform patient management, public health planning and practice, and disease prevention efforts for these populations.

## METHODS

Incidence data from 2003-2020 were obtained from USCS, which combines data from the NPCR (Centers for Disease Control and Prevention, CDC) and SEER programs. USCS includes all 50 states and the District of Columbia. This analysis covered 97% of the United States population as Nevada and Indiana were excluded due to incomplete data. Five-year survival data were collected from NPCR-funded registries from 2001-2019 and either had active case follow-up or were linked to the CDC’s National Death Index. NPCR covered 83% of the US population, excluding data from Connecticut, Hawaii, Iowa, Indiana, Massachusetts, Michigan, New Mexico, Nevada, South Dakota, Virginia, and Washington. Data from Kansas, Minnesota, North Dakota, and Wisconsin were excluded from the regression and survival curve analysis due to incomplete data. Unknown histological subtype or racial and ethnic group were additionally excluded. Cases were identified using International Classification of Disease for Oncology third edition (ICD-O-3) anatomy code C22.0 and histology codes 8170 (HCC not otherwise specified), 8171 (fibrolamellar), 8172 (scirrhous), 8173 (spindle cell variant), 8174 (clear cell type), and 8175 (pleomorphic type)^9^. Only primary tumors were included, and cases identified by autopsy or death certificate were excluded.

Data was stratified into ages 0-19 years old (pediatric cases: children and adolescents), and 20 years or older (adult cases), 0-14, 15-19, 20-29, 30-39, 40-64, and 65 years and older. Cases were stratified by sex, race and ethnicity, merged summary stage (local, regional, or distant)^10^, diagnosis year, metropolitan status by county, county-based economic status,^11^ and histology (fibrolamellar [8171] compared to all other HCC subtypes [8170, 8172-5]). Five-year relative survival (RS) was measured from 2001-2007 versus 2008-2019, corresponding to before and after Sorafenib approval for HCC in adults. For all analyses, statistics were not shown if a cell represented <6 cases.

Incidence was measured by counts and rates per 100,000 persons. Rates were age-adjusted using the 2000 United States standard population. Incidence trends were measured in annual percent change (APC) and average APC (AAPC) and calculated using Joinpoint software and defined as significant if different from zero using an alpha of 0.05. Trend analysis did not include the year 2020 due to data changes related to the COVID-19 pandemic^12^. Relative risk of incidence (RR) was estimated using negative binomial regression.

Five-year RS, defined as survival in the absence of death from other causes, was calculated via the complete method using expected life tables in SEER*Stat 8.4.2 (National Cancer Institute). RS results were considered different if 95% confidence intervals (CI) did not overlap. All-cause survival curves, overall and by demographic and clinical variables, were generated using the Kaplan–Meier method. Statistical testing for survival curves was performed using the log-rank test. Multivariable Cox proportional hazards modeling used separate models for pediatric and adult ages. Missing data were imputed (m=10 imputations) using the aregImpute function (Hmisc package in R). All predictor and outcome variables were included in the imputation process. Non-Hispanic American Indian/Alaska Native (NHAIAN) and Asian/Pacific Islander (NHAPI) patients were combined in regression analysis. The linearity assumption for continuous predictors was tested using restricted cubic spline functions. The proportional hazards (PH) assumption was assessed using the Schoenfeld residual correlation test. Schoenfeld residual plots were used to help determine time intervals within which the PH assumption holds. Histology violated the PH assumption in the model for children and adolescents. Therefore, a histology x time (<1 year vs. >1-5 years) interaction was included in the model to satisfy the PH assumption. Due to the large power to detect non-proportional hazards in the adult population, most variables violated the PH assumption. This violation was ignored so the hazard ratio (HR) presented among adults represent the average effect over five years of follow-up. Analysis was performed using SAS version 9.4 and R version 4.2.1.

## RESULTS

### Incidence of hepatocellular carcinoma

During 2003-2020, the pediatric incidence rate was 0.056 (95%CI:0.052-0.060; Table 1). In adults, the incidence rate was (7.793, 95%CI:7.767-7.819). Overall incidence was stable in the pediatric population at 0.3 average annual percent change (AAPC, 95%CI:-1.1-1.7) with no joinpoints identified (Fig.1A). In adults, AAPC was 2.9 (95%CI:2.8-3.1) driven by an initial increase from 2003-2009 (APC of 5.8) and 2009-2015 (3.0), and then declined during 2015-2019 (−1.5) (Fig. 1B).

**Figure 1.**
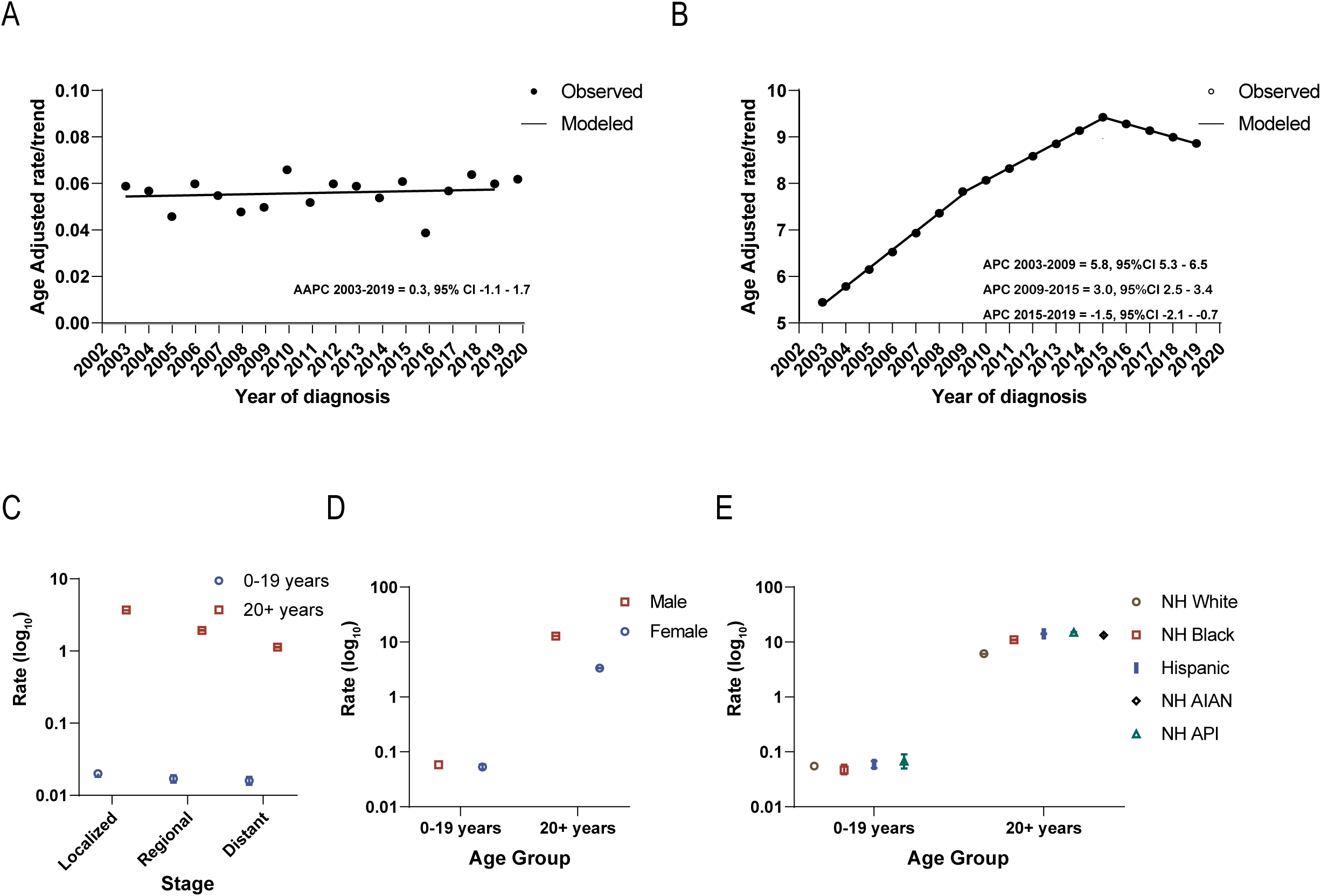
Incidence in hepatocellular carcinoma. A. Incidence trend of hepatocellular carcinoma cases in children and adolescents. B. Incidence trend of hepatocellular carcinoma cases in adults. C-E. Rate by stage. D. sex. E. race/ethnicity. Rates are per 100,000 persons. Abbreviations: annual percent change (APC), average annual percent change (AAPC), non-Hispanic white (NHW), Non-Hispanic Black (NHB), Non-Hispanic American Indian/Alaska Native (NHAIAN), Non-Hispanic Asian/Pacific Islander (NHAPI). Non-Hispanic American Indian/Alaska Native were excluded in pediatric ages due to <6 cases. Incidence trends were measured in APC and AAPC and calculated using Joinpoint software and defined as significant if different from zero using an alpha of 0.05. Error bars indicate confidence intervals.

**Table 1.** Incidence of hepatocellular carcinoma cases, with multivariable negative binomial analysis, United States Cancer Statistics database, 2003-2020.

| Variable | 0-19 years |  |  |  | 20+ years |  |  |  |
| --- | --- | --- | --- | --- | --- | --- | --- | --- |
|  | Count | Rate (95%CI) | RR (95%CI) | p-value | Count | Rate (95%CI) | RR (95%CI) | p-value |
| Total | 813 | 0.056 (0.052-0.060) | ~ | ~ | 354,536 | 7.793 (7.767-7.819) | ~ | ~ |
| Sex |  |  |  |  |  |  |  |  |
| Male | 431 | 0.058 (0.053-0.064) | ref | ref | 273,079 | 12.891 (12.842-12.941) | ref | ref |
| Female | 382 | 0.054 (0.048-0.059) | 0.93 (0.81-1.07) | 0.31 | 81,457 | 3.363 (3.339-3.386) | 0.30 (0.29-0.32) | <0.0001 |
| Race and Ethnicity |  |  |  |  |  |  |  |  |
| NHW | 444 | 0.056 (0.051-0.061) | ref | ref | 205,091 | 6.150 (6.123-6.177) | ref | ref |
| NHB | 106 | 0.048 (0.039-0.058) | 0.84 (0.67-1.05) | 0.12 | 52,670 | 10.819 (10.724-10.914) | 1.92 (1.79-2.06) | <0.0001 |
| NHAIAN | ~ | ~ | 0.69 (0.28-1.67) | 0.41 | 3,917 | 13.439 (13.002-13.886) | 2.21 (2.02-2.40) | <0.0001 |
| NHAPI | 52 | 0.069 (0.051-0.090) | 1.12 (0.83-1.51) | 0.46 | 28,918 | 14.706 (14.533-14.881) | 2.38 (2.21-2.57) | <0.0001 |
| Hispanic | 185 | 0.058 (0.050-0.067) | 1.02 (0.85-1.22) | 0.86 | 56,815 | 14.042 (13.922-14.162) | 1.83 (1.71-1.96) | <0.0001 |
| Stage |  |  |  |  |  |  |  |  |
| Localized | 299 | 0.021 (0.018-0.023) | ~ | ~ | 168,952 | 3.707 (3.690-3.725) | ~ | ~ |
| Regional | 247 | 0.017 (0.015-0.019) | ~ | ~ | 88,873 | 1.948 (1.935-1.961) | ~ | ~ |
| Distant | 234 | 0.016 (0.014-0.018) | ~ | ~ | 51,587 | 1.137 (1.127-1.147) | ~ | ~ |
| Histology |  |  |  |  |  |  |  |  |
| HCC (non-fHCC) | 492 | 0.034 (0.031-0.037) | ~ | ~ | 353,648 | 7.772 (7.746-7.798) | ~ | ~ |
| fHCC | 321 | 0.022 (0.020-0.024) | ~ | ~ | 888 | 0.021 (0.020-0.023) | ~ | ~ |

8 Rates per 100,000 persons. Three decimal places were used for pediatric ages instead of expressing rates per 1 million. Abbreviations: fibrolamellar hepatocellular 9 carcinoma (fHCC), hepatocarcinoma (HCC), Non-Hispanic white (NHW), Non-Hispanic Black (NHB), Non-Hispanic American Indian/Alaska Native (NHAIAN), Non-Hispanic Asian and Pacific Islanders (NHAPI), relative risk (RR), confidence intervals (CI), reference group (ref), too few cases to calculate (~).

In adults, the incidence of localized (3.707, 95%CI:3.690-3.725) was higher than regional (1.948, 95%CI:1.935-1.961) or distant disease (1.137, 95%CI:1.127-1.147). Among pediatric cases, incidence was similar for localized (0.021, 95%CI:0.018-0.023), regional (0.017, 95%CI:0.015-0.019), and distant disease (0.016, 95%CI:0.014-0.018) (Fig.1E).

There was no overall difference in incidence between pediatric (0.022, 95%CI:0.020-0.024) and adult (0.021, 95%CI:0.020-0.023) fibrolamellar HCC (fHCC) (Table 1). The incidence of fHCC was less than that of other HCC types in all age groups, except for adolescents (15-19 years) who had the highest incidence (0.046, 95%CI:0.059-0.064) (eTable 1).

In children and adolescents, HCC incidence was similar between males (0.058, 95%CI:0.053-0.064) and females (0.054, 95%CI:0.048-0.059). In contrast, in adults, males had a higher incidence of HCC (12.891, 95%CI:12.842-12.941) compared to females (3.363, 95%CI:3.339-3.386) (Fig.1D). For pediatric HCC, incidence was similar regardless of race and ethnicity, while it varied in adults (Fig. 1 E).

Relative risk (RR) of developing HCC was analyzed in pediatric and adult populations. Risk increased with age in both pediatric and adult groups. Individuals aged 15-19 years (p<0.001) had higher risk compared to those aged 0-14 years (3.15 95%CI:2.73-3.64), and risk was higher (p<0.001) for all older age groups in adults compared to individuals in the 20–29-year group (eTable 2). In adults, racial and ethnic group, socioeconomic status, and metropolitan county size, were risk factors, but were similar for pediatric ages.

### Five-year relative survival of patients with hepatocellular carcinoma

Pediatric 5-year RS was 46.4% (95%CI:42.4-50.3) and was 20.7% (95%CI:20.5-20.9; Table 2) in adults. Survival was lower with increasing disease stage for both pediatric and adult populations. Pediatric cases had better outcomes compared to adults for all stages: localized disease (75.1%, 95%CI:68.8-80.4 versus 33.6%, 95%CI:33.3-34.0), regional (44.5%, 95%CI:37.1-51.6 versus 12.0% 11.7-12.2), and distant disease (14.1%, 95%CI:9.5-19.7 versus 3.5%, 95%CI:3.2-3.7), in children versus adults, respectively (Fig.2A).

**Figure 2.**
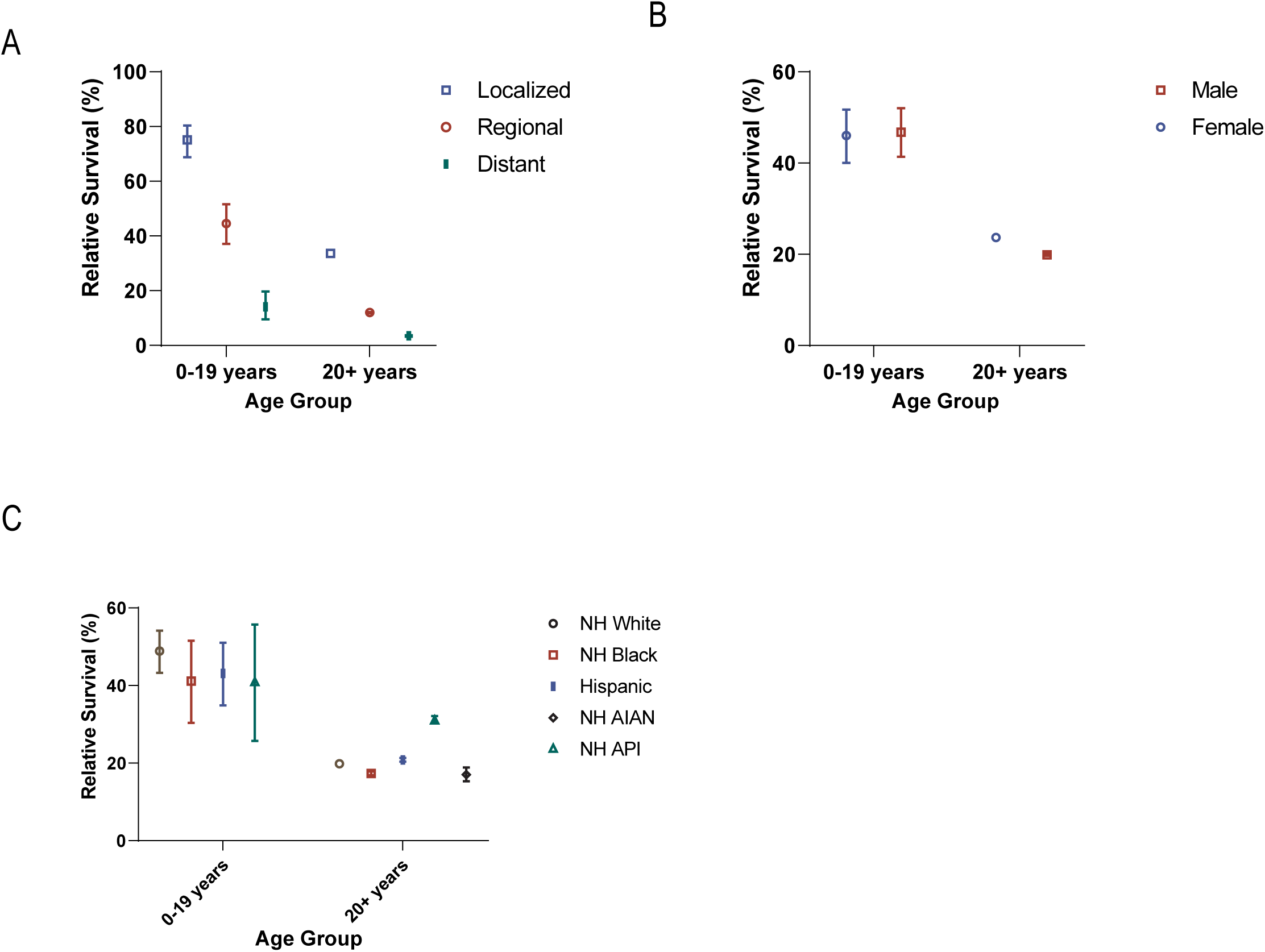
5-year relative survival of patients with hepatocellular carcinoma. A. stage. B. sex. C. race/ethnicity. Abbreviations: Non-Hispanic (NH), American Indian/Alaskan Native (NHAIAN), Asian/Pacific Islander (NHAPI), annual percent change (APC). Non-Hispanic American Indian/Alaska Native were excluded in pediatric due to <6 cases. Error bars indicate confidence intervals.

**Table 2.**
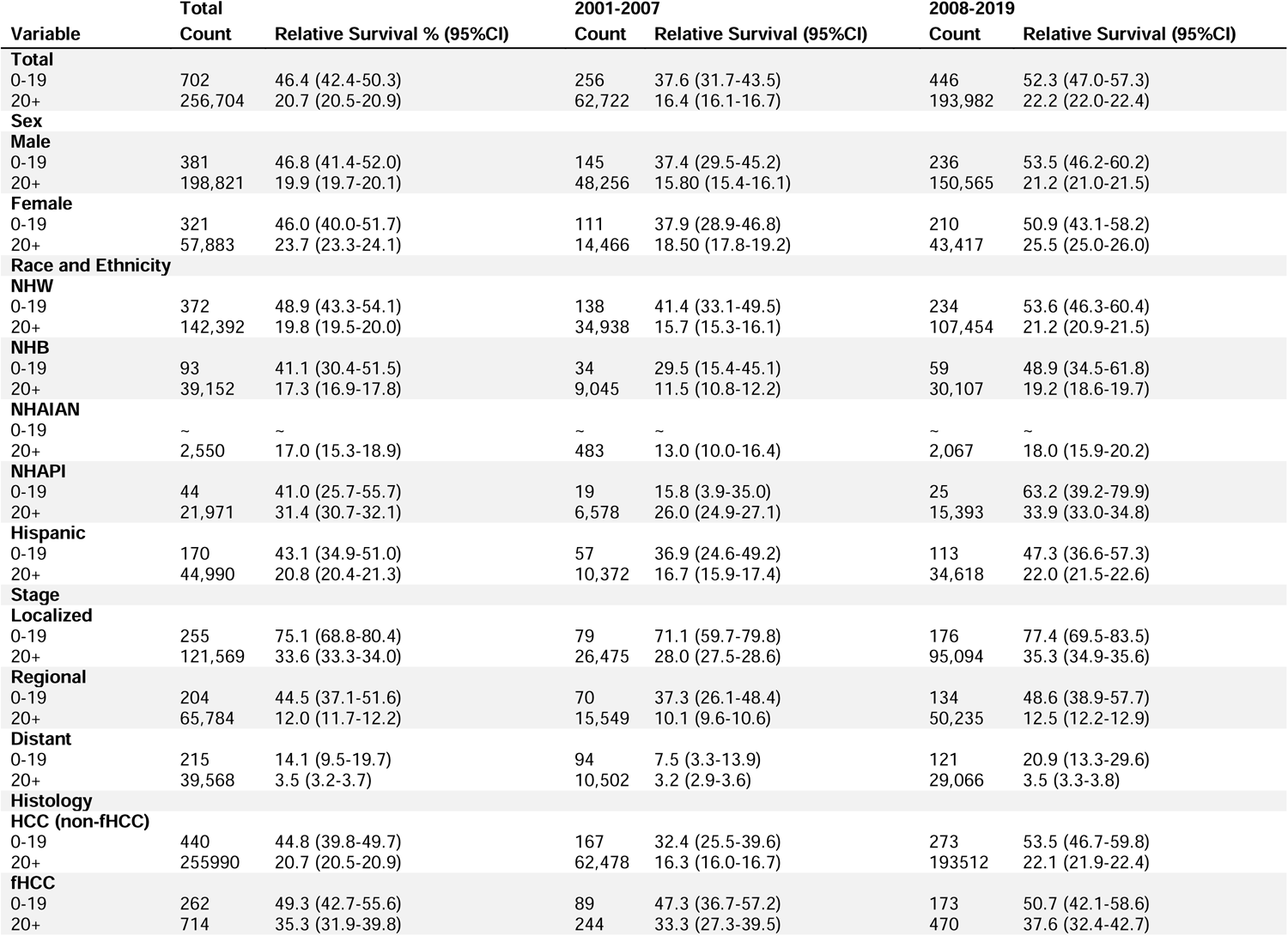
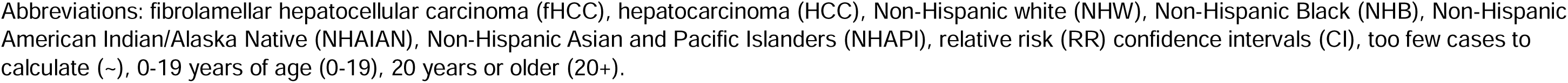
Relative survival of cases with hepatocellular carcinoma, National Program of Cancer Registries, 2001-2019.

| Variable | Total | Relative Survival % (95%CI) | 2001-2007 |  | 2008-2019 |  |
| --- | --- | --- | --- | --- | --- | --- |
|  | Count |  | Count | Relative Survival (95%CI) | Count | Relative Survival (95%CI) |
| **Total** |  |  |  |  |  |  |
| 0-19 | 702 | 46.4 (42.4-50.3) | 256 | 37.6 (31.7-43.5) | 446 | 52.3 (47.0-57.3) |
| 20+ | 256,704 | 20.7 (20.5-20.9) | 62,722 | 16.4 (16.1-16.7) | 193,982 | 22.2 (22.0-22.4) |
| **Sex** |  |  |  |  |  |  |
| **Male** |  |  |  |  |  |  |
| 0-19 | 381 | 46.8 (41.4-52.0) | 145 | 37.4 (29.5-45.2) | 236 | 53.5 (46.2-60.2) |
| 20+ | 198,821 | 19.9 (19.7-20.1) | 48,256 | 15.80 (15.4-16.1) | 150,565 | 21.2 (21.0-21.5) |
| **Female** |  |  |  |  |  |  |
| 0-19 | 321 | 46.0 (40.0-51.7) | 111 | 37.9 (28.9-46.8) | 210 | 50.9 (43.1-58.2) |
| 20+ | 57,883 | 23.7 (23.3-24.1) | 14,466 | 18.50 (17.8-19.2) | 43,417 | 25.5 (25.0-26.0) |
| **Race and Ethnicity** |  |  |  |  |  |  |
| **NHW** |  |  |  |  |  |  |
| 0-19 | 372 | 48.9 (43.3-54.1) | 138 | 41.4 (33.1-49.5) | 234 | 53.6 (46.3-60.4) |
| 20+ | 142,392 | 19.8 (19.5-20.0) | 34,938 | 15.7 (15.3-16.1) | 107,454 | 21.2 (20.9-21.5) |
| **NHB** |  |  |  |  |  |  |
| 0-19 | 93 | 41.1 (30.4-51.5) | 34 | 29.5 (15.4-45.1) | 59 | 48.9 (34.5-61.8) |
| 20+ | 39,152 | 17.3 (16.9-17.8) | 9,045 | 11.5 (10.8-12.2) | 30,107 | 19.2 (18.6-19.7) |
| **NHAIAN** |  |  |  |  |  |  |
| 0-19 | ~ | ~ | ~ | ~ | ~ | ~ |
| 20+ | 2,550 | 17.0 (15.3-18.9) | 483 | 13.0 (10.0-16.4) | 2,067 | 18.0 (15.9-20.2) |
| **NHAPI** |  |  |  |  |  |  |
| 0-19 | 44 | 41.0 (25.7-55.7) | 19 | 15.8 (3.9-35.0) | 25 | 63.2 (39.2-79.9) |
| 20+ | 21,971 | 31.4 (30.7-32.1) | 6,578 | 26.0 (24.9-27.1) | 15,393 | 33.9 (33.0-34.8) |
| **Hispanic** |  |  |  |  |  |  |
| 0-19 | 170 | 43.1 (34.9-51.0) | 57 | 36.9 (24.6-49.2) | 113 | 47.3 (36.6-57.3) |
| 20+ | 44,990 | 20.8 (20.4-21.3) | 10,372 | 16.7 (15.9-17.4) | 34,618 | 22.0 (21.5-22.6) |
| **Stage** |  |  |  |  |  |  |
| **Localized** |  |  |  |  |  |  |
| 0-19 | 255 | 75.1 (68.8-80.4) | 79 | 71.1 (59.7-79.8) | 176 | 77.4 (69.5-83.5) |
| 20+ | 121,569 | 33.6 (33.3-34.0) | 26,475 | 28.0 (27.5-28.6) | 95,094 | 35.3 (34.9-35.6) |
| **Regional** |  |  |  |  |  |  |
| 0-19 | 204 | 44.5 (37.1-51.6) | 70 | 37.3 (26.1-48.4) | 134 | 48.6 (38.9-57.7) |
| 20+ | 65,784 | 12.0 (11.7-12.2) | 15,549 | 10.1 (9.6-10.6) | 50,235 | 12.5 (12.2-12.9) |
| **Distant** |  |  |  |  |  |  |
| 0-19 | 215 | 14.1 (9.5-19.7) | 94 | 7.5 (3.3-13.9) | 121 | 20.9 (13.3-29.6) |
| 20+ | 39,568 | 3.5 (3.2-3.7) | 10,502 | 3.2 (2.9-3.6) | 29,066 | 3.5 (3.3-3.8) |
| **Histology** |  |  |  |  |  |  |
| **HCC (non-fHCC)** |  |  |  |  |  |  |
| 0-19 | 440 | 44.8 (39.8-49.7) | 167 | 32.4 (25.5-39.6) | 273 | 53.5 (46.7-59.8) |
| 20+ | 255990 | 20.7 (20.5-20.9) | 62,478 | 16.3 (16.0-16.7) | 193512 | 22.1 (21.9-22.4) |
| **fHCC** |  |  |  |  |  |  |
| 0-19 | 262 | 49.3 (42.7-55.6) | 89 | 47.3 (36.7-57.2) | 173 | 50.7 (42.1-58.6) |
| 20+ | 714 | 35.3 (31.9-39.8) | 244 | 33.3 (27.3-39.5) | 470 | 37.6 (32.4-42.7) |

Abbreviations: fibrolamellar hepatocellular carcinoma (fHCC), hepatocarcinoma (HCC), Non-Hispanic white (NHW), Non-Hispanic Black (NHB), Non-Hispanic American Indian/Alaska Native (NHAIAN), Non-Hispanic Asian and Pacific Islanders (NHAPI), relative risk (RR) confidence intervals (CI), too few cases to calculate (~), 0-19 years of age (0-19), 20 years or older (20+).

In adults, 5-year RS for with fHCC (35.3%, 95%CI:31.9-39.8) was better than for those with other HCC subtypes (20.7%, 95%CI:20.5-20.9), but these measures were similar in pediatric cases (49.3%, 95%CI:42.7-55.6 versus 44.8%, 95%CI:39.8-49.7, respectively). Unlike other HCC types, survival reported for fHCC did not improve between 2001 and 2007 versus 2008 and 2019 for either pediatric (47.3%, 95%CI:36.7-57.2 versus 50.7%, 95%CI:42.1-58.6) or adult populations (33.3%, 95%CI:27.3-39.5 versus 37.6%, 95%CI:32.4-42.7).

Over time, in adults, RS increased overall from 16.4% (95%CI:16.1-16.7) between 2001 and 2007 to 22.2% (95%CI:22.0-22.4) between 2008 and 2019. In pediatric ages during the same periods, RS increased from 37.6% (95%CI:31.7-43.5) to 52.3% (95%CI:47.0-57.3).

Pediatric ages had similar RS between males (46.8%, 95%CI:41.4-52.0) and females (46.0%, 95%CI:40.0-51.7). In adults, RS was lower in males (19.9%, 95%CI:19.7-20.1) than females (23.7, 95%CI:23.3-24.1) (Fig.2B). RS was also similar in pediatric ages regardless of race and ethnicity but varied in adults (Fig.2C).

### All-cause survival analysis

In children and adolescents, the risk of death within five years was significantly lower for cases diagnosed in 2008 or later compared to those diagnosed from 2001 to 2007 (HR =0.69, 95%CI:0.55-0.86). Compared to local disease, regional (HR=3.10, 95%CI:2.22-4.32), and distant (HR=7.18, 95%CI:5.25-9.82) disease stages were associated with a higher risk of death. Histology was also a significant predictor of 5-year survival (p<0.0001), although this relationship varied over time. Fibrolamellar HCC was associated with a lower risk of death within the first year of follow-up compared to other HCC histology types (HR=0.38, 95%CI:0.26-0.55) but after one year, the risk of death within five years for fHCC vs other HCC types was similar (HR=1.06, 95%CI:0.77-1.48). Non-Hispanic Black race and ethnicity showed higher risk of death compared to non-Hispanic White race and ethnicity (HR=1.48, 95%CI:1.07-2.05). Children and adolescents diagnosed in counties with a metropolitan population of 250,000–1 million (HR=1.36, 95%CI:1.02-1.82) and non-metropolitan areas (HR=1.45, 95%CI:1.01-2.08) had a higher risk of death compared to those diagnosed in metropolitan counties with a population >1 million. Age, sex, and county economic status were not significant predictors of five-year survival (Fig.3A, eFig.1, eFig.2).

**Figure 3.**
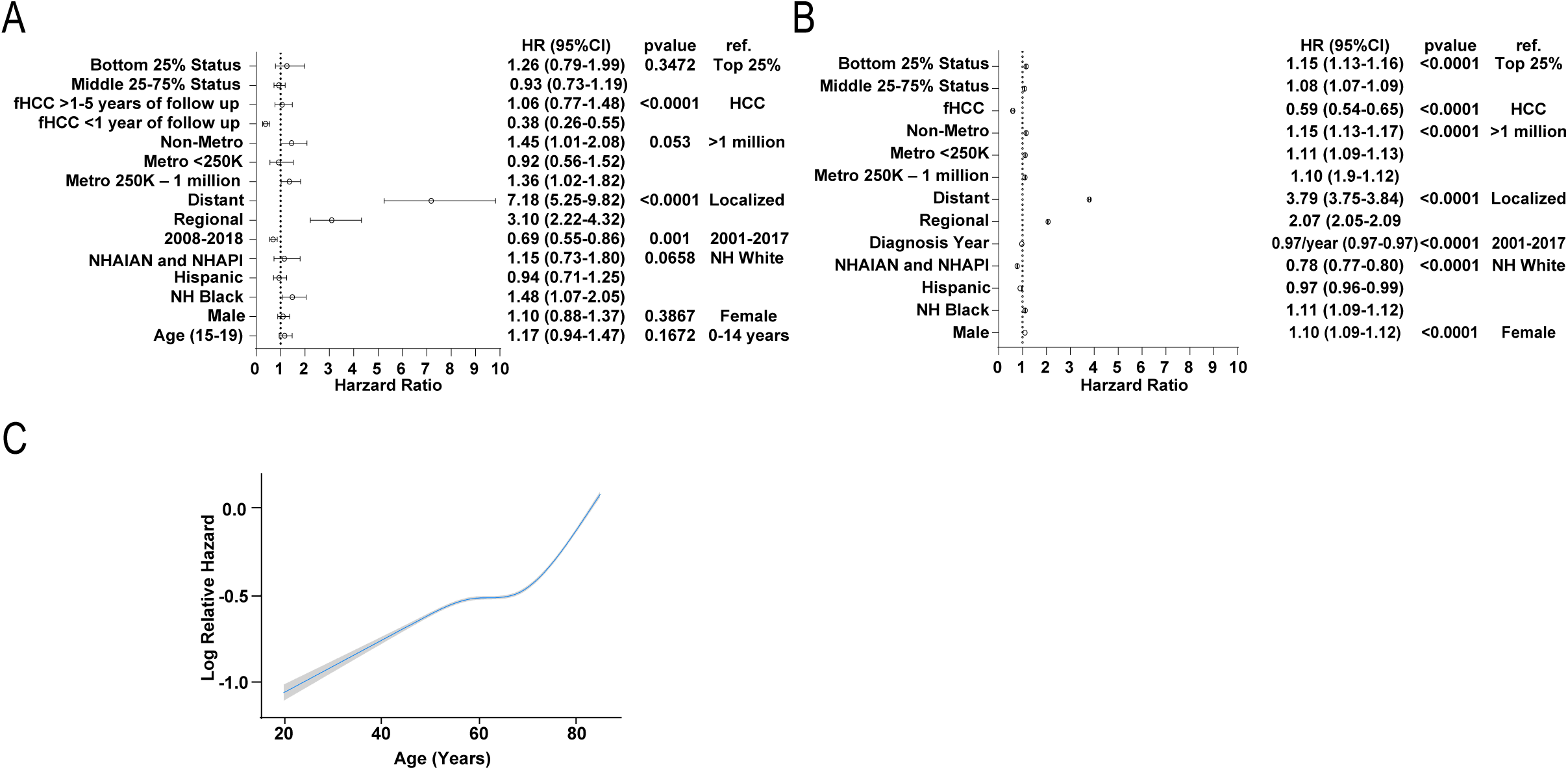
Overall survival of patients with hepatocellular carcinoma. A-B 5-year overall survival of patients with hepatocellular carcinoma. A. Pediatric risk factors. B. Adult risk factors C. Modeling of relative hazard against age in adults. Abbreviations: hazard ratio (HR), reference group (ref.), 95% confidence interval (CI), metropolitan (metro), Non-Hispanic American Indian/Alaska Native (NHAIAN) or Non-Hispanic Asian/Pacific Islander (NHAPI). Error bars indicate confidence intervals. P-value was calculated from multivariate Cox analysis for each group; individual subgroups were considered significant if CI did not cross 1 (p<.05).

In adults, we assessed later diagnosis year per 1-year increase as the data was linear (HR=0.97, 95%CI:0.97-0.97). Compared to adults with local disease, regional (HR-2.07, 95%CI:2.05-2.09) and distant disease (HR=3.79, 95%CI:3.75-3.84) were also associated higher risk of death. Fibrolamellar HCC (HR=0.59, 95%CI:0.54-0.65) was associated with a lower risk of death compared to other histological types of HCC. Male sex (HR=1.10, 95%CI:1.09-1.12), non-Hispanic Black race and ethnicity (HR=1.11, 95%CI:1.09-1.12 compared to non-Hispanic White), bottom 25% (HR=1.15, 95%CI:1.13-1.16 compared to top 25%) and 25%-75% county economic status (HR=1.08, 95%CI:1.07-1.09), and lower county population were all associated with higher risk of death (Fig. 3B, eFig.1, eFig.2). Age was associated with higher risk of death although the relationship was non-linear (Fig.3C).

## DISCUSSION

Using data from high coverage databases, we described the incidence and survival of children, adolescents, and adults with HCC. We show that the recent decline in HCC incidence described in adults has not occurred in children and adolescents with HCC, extending findings from smaller studies and suggesting that incidence in children has been stable since 1973^2,3,13–17^. It is possible that the etiological shift from HCV- to MASLD -driven disease that has been proposed to explain this trend in adults may not apply to children^13–17^. Pediatric MASLD incidence is increasing in the United States, and it is unclear if the increase in this risk factor would lead to an increase in HCC in children. Our data demonstrates unchanged incidence of HCC despite the increase in pediatric MASLD in the US^13–16,18,19^.

We show that among pediatric ages, most are diagnosed with local disease as opposed to regional or distant disease, clarifying conflicting results from prior smaller studies^2–4^. Moreover, consistent with a prior study, we show that children and adolescents are more frequently diagnosed with advanced disease than adults^4^. This finding likely reflects the higher proportion of *de novo* HCC in children, as these cases are diagnosed at more advanced disease stage while adults are surveilled after onset of cirrhosis^20^.

Our results confirm the well-established higher incidence of HCC in adult males versus females^2–5,13,14^. In contrast to most studies in the pediatric population, we found no difference in HCC incidence based on sex^2–5^. Recently, it was shown that the male-to-female ratio declined in adults aged <50 years from 2009-2015, with multiple studies showing a faster decline in incidence among males than females in recent years^13–15^. This shift from a predominantly male disease has been linked by age-period-cohort analysis to recent shifts in etiology^13^.

We found no increased risk of developing HCC based on demographic factors in children and adolescents. In contrast, in adults there were clear disparities based on race and ethnicity, metropolitan status, and socioeconomic status highlighting the need to incorporate demographic factors in the prevention and treatment of HCC in adults. This difference is likely due to the different etiologies in children versus adults^2,3,5,8,13–17^. Consistent with the adult literature, we show an increase in risk with age, which in children likely reflects the higher incidence of fHCC in adolescents^2–4,13^.

Interestingly, risk of death from HCC did not increase linearly with age, instead showing a plateau before a sharp increase in older ages. Although younger adults more frequently present with less favorable tumor characteristics, preserved liver function in younger individuals, more aggressive therapy, and better post-operative recovery may all contribute to this effect^21–23^.

Survival was higher in pediatric ages compared to adults regardless of disease stage, however pediatric ages had a higher proportion of advanced disease overall. Surgery remains the cornerstone of treatment for both pediatric and adult HCC^2,4,24^. A higher proportion of pediatric patients undergo surgery than adults, possibly due to the lower prevalence of underlying liver disease; this likely contributes to better survival for pediatric patients^4^. Multivariate analysis showed that survival improved in all groups over time, with risk of death decreasing linearly with later year of diagnosis in adults. Both children and adults had better survival in 2008-2019 versus 2001-2007. In pediatric cases, survival improved over time in all stages, but confidence intervals overlapped.

Consistent with other reports, we found that adult females with HCC survived longer than males. This is consistent with the idea that sex hormones may play a role in improving the survival of women with HCC^8^. In children, however, we found no difference between the sexes while previous reports present mixed findings^2,3,8,24^.

We found a 48% higher risk of death at 5 years for non-Hispanic Black compared to non-Hispanic White children and adolescents and an 11% higher risk for non-Hispanic Black compared to non-Hispanic White adults. This effect was apparent after controlling for socio-economic status and disease stage, which are known to influence racial and ethnic disparities^8,25^. Moreover, we did not find a significant difference in survival among pediatric ages according to socioeconomic status that was described in the adult population. Collectively, this disparity may be mediated by known differences in quality of care, and poor provider-patient interactions including bias or distrust.^8,24,26,27^. Kahla et al. recently published a similar finding in hepatoblastoma^28^. This may point to broader inequity in the treatment of liver disease that may be driven by unequal transplant care, particularly living donor transplant and waitlist mortality that are partially independent of economic status^29,30^. We also found lower survival in smaller metropolitan counties, similar to what has been described for adults with HCC, which may reflect decreased quality of care^31^.

Consistent with previous population-based studies, we found that fHCC was associated with better overall survival compared to all other HCC histological types. Prior studies attributed this difference to more aggressive surgical treatment, less frequent underlying liver disease, less aggressive biology, and younger age of patients with fHCC^2–4,32–34^. This finding remains controversial as the International Childhood Liver Tumors Strategy Group (SIOPEL) reported no difference in three-year follow-up between fHCC and HCC but better one-year survival in HCC^35^. Our study reconciles the results from SIOPEL and those of population-based studies. We found lower risk in pediatric ages within one year of follow-up, but no difference with longer follow up times, possibly due to higher complete resection rate or delayed follow-up in fHCC^32–35^. While adults with fHCC had better overall survival than adults with other HCCs, the survival benefit persists beyond one year, possibly due to lower resection rates and higher frequency of underlying liver disease in adults compared to children^32–35^. Importantly, we found that fHCC survival did not improve in the 2008-2019 period compared to 2001-2017 unlike other HCC type survival, suggesting that fHCC-specific treatments may be needed to improve outcomes.

We acknowledge several limitations of our study. First, there were insufficient numbers of NHAIAN or NHAPI cases to consider these groups separately in our survival analyses. We may be underpowered to detect small differences in incidence based on race and ethnicity. Additionally, due to significant heterogeneity and large sample size in the adult population, we were unable to satisfy the proportional hazards assumption for the multivariate survival analysis. We acknowledge that county-wide measures of socioeconomic status and population may not capture significant heterogeneity within each county. Our study is limited to the information contained in the USCS and NPCR databases and provides no validated information relating to etiology, tumor characteristics beyond stage, or treatment such as surgery.

In summary, the incidence of pediatric HCC remained stable between 2003-2019, unlike the adult population that experienced a recent decrease. We show that several of the demographic disparities found in adults do not extend to children and adolescents, including the higher incidence in males and in racial and ethnic groups other than non-Hispanic White, as well as better survival in females. We describe several novel differences in outcomes including a higher risk of death for non-Hispanic Black children, adolescents, and adults; additional research and potential interventions may improve outcomes in this population^36^. We are also the first to report on metropolitan and area-level socioeconomic status in pediatric HCC, showing a similar higher in risk of death for both pediatric and adult ages associated with decreasing county size; future research may be helpful to better understand this pattern for pediatric ages. Lastly, this is the first population-based study to confirm that compared to other HCC types, the previously described survival benefit in children with fHCC is limited to short-term follow up and does not translate to long term survival. While there are similarities between pediatric and adult HCC, there are several important differences that highlight the need for age-specific (pediatric vs adult) research and risk-adapted management strategies that incorporate the demographic vulnerabilities defined here for patients with HCC.

## Data Availability

The data that support the findings of this study are available on request
by contacting. The data are not publicly available
due to privacy and legal restrictions.

## ACKNOWLEDGEMENTS

The authors would like to thank Dr. Hashem B. El-Serag for his helpful discussion regarding the preliminary analyses.

## FUNDING

This work was supported by the National Institute of General Medical Sciences of the National Institutes of Health under Award Number T32GM136554, and internal funding from the Center from Advanced Innate Cell Therapy of Texas Children’s Cancer Center (AH).

## AUTHORS CONTRIBUTIONS

AA Conceptualization, Writing-Original Draft, Visualization, Project Administration, Funding Acquisition. DAS Conceptualization, Writing-Review and Editing, Methodology, Software, Formal analysis. SD, Methodology, Software, Formal analysis. TDT Writing-Review and Editing, Methodology, Software, Formal analysis. JF Writing-Review and Editing, EJP Writing-Review and Editing, BM Writing-Review and Editing, PJL Writing-Review and Editing, AH Conceptualization, Writing-Review and Editing, Project Administration, Funding Acquisition.

## COMPETING INTERESTS STATEMENT

Andras Heczey has consultancy / scientific advisory roles with Waypoint Bio and Cargo Therapeutics. All the other authors declare that they have no competing financial interests.

---

**eFigure 1.**
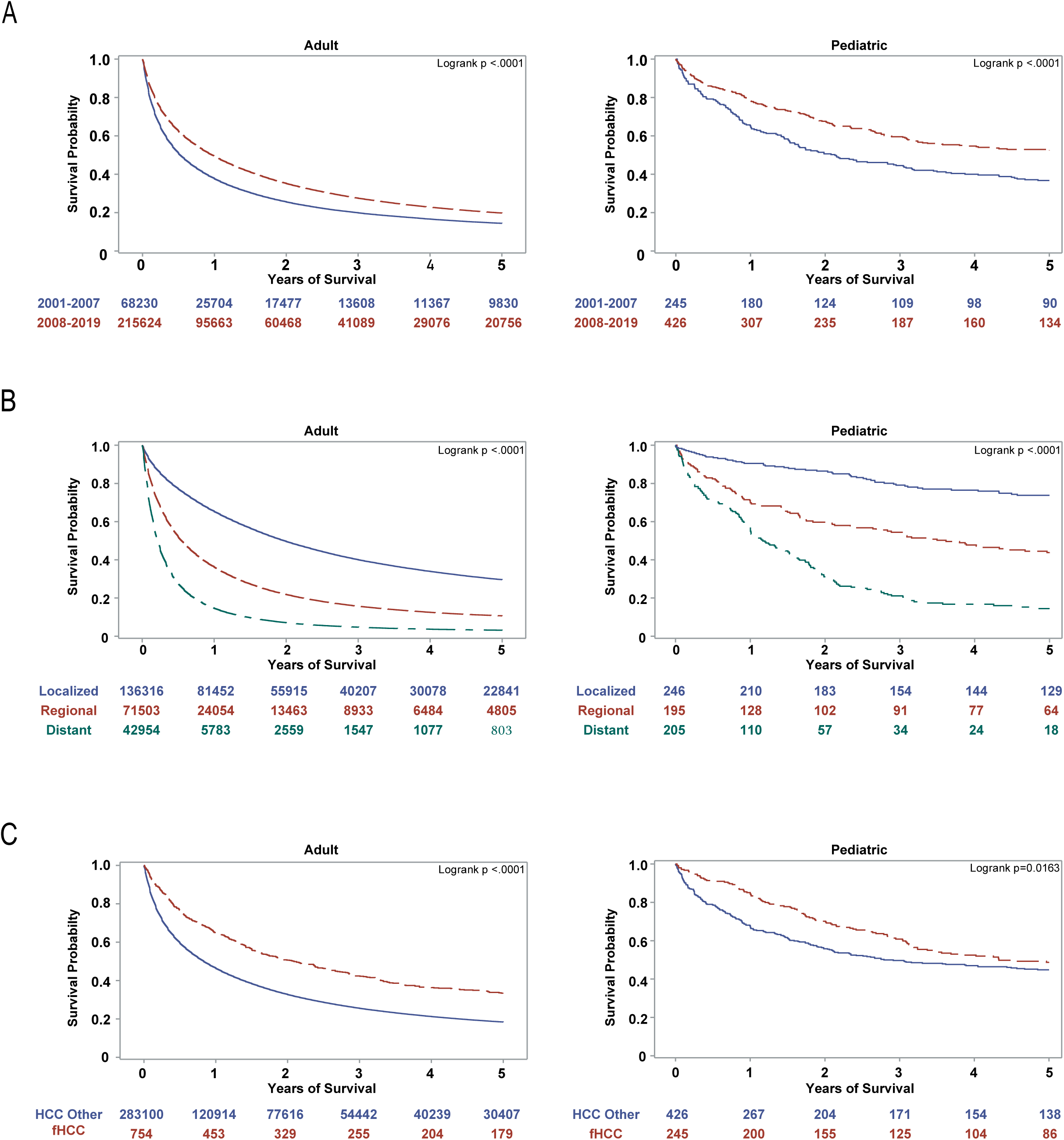
5-year overall survival of adult (left) and pediatric (right) patients with hepatocellular carcinoma. A. Diagnosis year. B. Stage. C. Histology. Abbreviation: fibrolamellar hepatocellular carcinoma (fHCC), hepatocarcinoma (HCC).

**eFigure 2.**
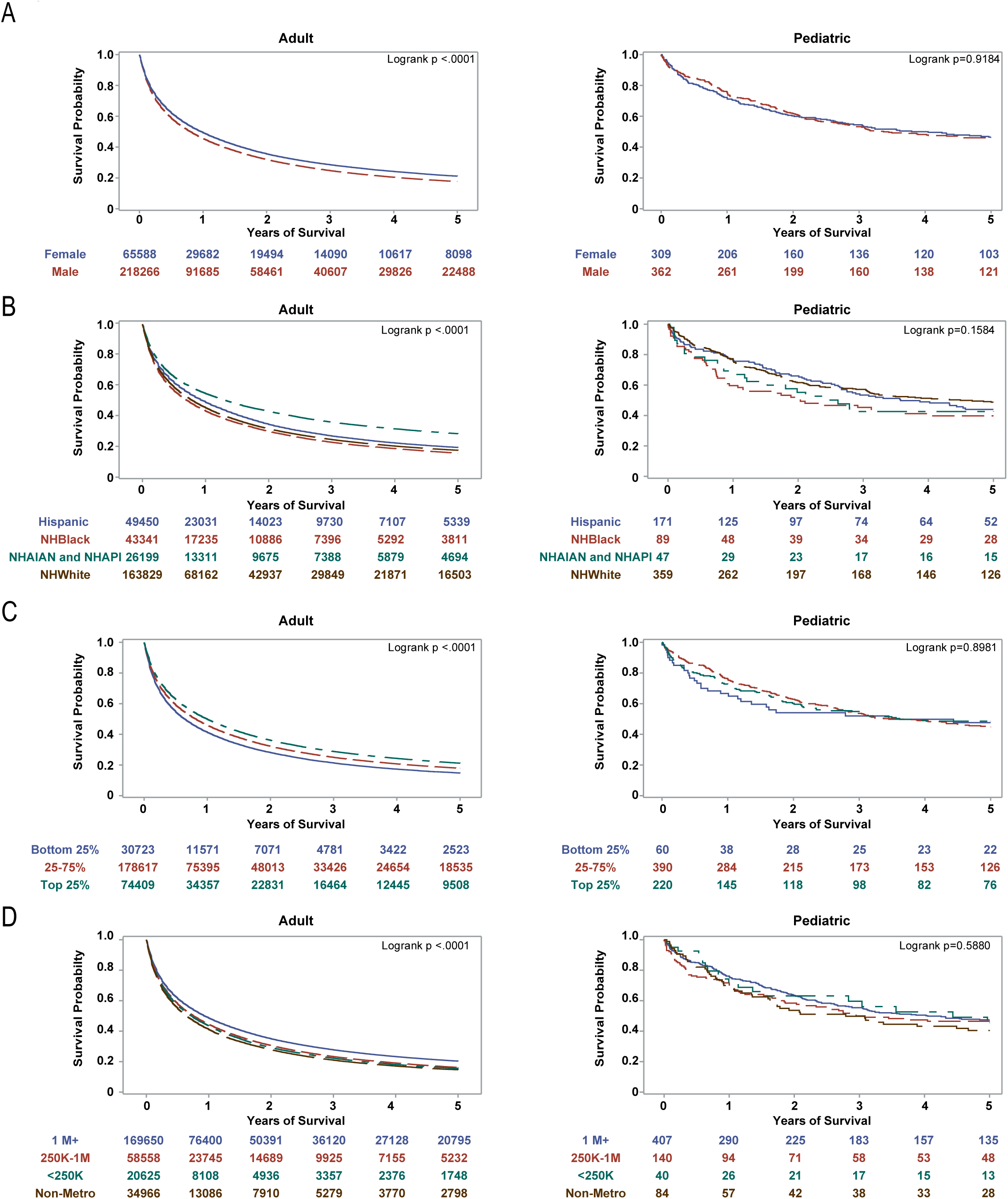
5-year overall survival of adult (left) and pediatric (right) patients with hepatocellular carcinoma. A. Sex. B. Race and ethnicity. C. Socioeconomic status by county. D. Metropolitan status. Abbreviation: metropolitan (metro), Non-Hispanic (NH).

## Notes

### Funding Statement

Research reported in this publication was supported by the National Institute of General Medical Sciences of the National Institutes of Health under Award Number T32GM136554 and internal funding from the Center for Advanced Innate Cell Therapy of Texas Childrens Cancer Center Houston United States.

### Summary of Updates

Author names adjusted. Disclosure of advisory role added.

## REFERENCES

1. Rumgay H, Arnold M, Ferlay J, et al. Global burden of primary liver cancer in 2020 and predictions to 2040. J Hepatol. 2022;77(6):1598–1606. doi:10.1016/j.jhep.2022.08.021

2. McAteer JP, Goldin AB, Healey PJ, Gow KW. Hepatocellular carcinoma in children: epidemiology and the impact of regional lymphadenectomy on surgical outcomes. J Pediatr Surg. 2013;48(11):2194–2201. doi:10.1016/j.jpedsurg.2013.05.007

3. Allan BJ, Wang B, Davis JS, et al. A review of 218 pediatric cases of hepatocellular carcinoma. J Pediatr Surg. 2014;49(1):166–171; discussion 171. doi:10.1016/j.jpedsurg.2013.09.050

4. Lau CSM, Mahendraraj K, Chamberlain RS. Hepatocellular Carcinoma in the Pediatric Population: A Population Based Clinical Outcomes Study Involving 257 Patients from the Surveillance, Epidemiology, and End Result (SEER) Database (1973-2011). HPB Surg. 2015;2015:670728. doi:10.1155/2015/670728

5. Khanna R, Verma SK. Pediatric hepatocellular carcinoma. World J Gastroenterol. 2018;24(35):3980–3999. doi:10.3748/wjg.v24.i35.3980

6. Czauderna P. Adult type vs. Childhood hepatocellular carcinoma--are they the same or different lesions? Biology, natural history, prognosis, and treatment. Med Pediatr Oncol. 2002;39(5):519–523. doi:10.1002/mpo.10178

7. Weeda VB, Aronson DC, Verheij J, Lamers WH. Is hepatocellular carcinoma the same disease in children and adults? Comparison of histology, molecular background, and treatment in pediatric and adult patients. Pediatric Blood & Cancer. 2019;66(2):e27475. doi:10.1002/pbc.27475

8. Guo A, Pomenti S, Wattacheril J. Health Disparities in Screening, Diagnosis, and Treatment of Hepatocellular Carcinoma. Clin Liver Dis (Hoboken). 2021;17(5):353–358. doi:10.1002/cld.1057

9. World Health Organization. International Classification of Diseases for Oncology (ICD-O). 3rd ed., 1st revision. World Health Organization; 2013. Accessed March 15, 2024. https://iris.who.int/handle/10665/96612

10. Review of Staging Systems | SEER Training. Accessed March 15, 2024. https://training.seer.cancer.gov/collaborative/intro/systems_review.html

11. Distressed Designation and County Economic Status Classification System. Appalachian Regional Commission. Accessed December 1, 2023. https://www.arc.gov/distressed-designation-and-county-economic-status-classification-system/

12. Mariotto AB, Feuer EJ, Howlader N, Chen HS, Negoita S, Cronin KA. Interpreting cancer incidence trends: challenges due to the COVID-19 pandemic. JNCI: Journal of the National Cancer Institute. 2023;115(9):1109–1111. doi:10.1093/jnci/djad086

13. Zhang X, El-Serag HB, Thrift AP. Sex and Race Disparities in the Incidence of Hepatocellular Carcinoma in the United States Examined through Age–Period–Cohort Analysis. *Cancer Epidemiology*, Biomarkers & Prevention. 2020;29(1):88–94. doi:10.1158/1055-9965.EPI-19-1052

14. Rich NE, Yopp AC, Singal AG, Murphy CC. Hepatocellular Carcinoma Incidence is Decreasing Among Younger Adults in the United States. Clin Gastroenterol Hepatol. 2020;18(1):242–248.e5. doi:10.1016/j.cgh.2019.04.043

15. Han J, Wang B, Liu W, et al. Declining disease burden of HCC in the United States, 1992–2017: A population-based analysis. Hepatology. 2022;76(3):576. doi:10.1002/hep.32355

16. Shiels MS, O’Brien TR. Declining U.S. Hepatocellular Carcinoma Rates, 2014–2017. Clin Gastroenterol Hepatol. 2022;20(2):e330–e334. doi:10.1016/j.cgh.2021.02.011

17. Thrift AP, Liu KS, Raza SA, El-Serag HB. Recent Decline in the Incidence of Hepatocellular Carcinoma in the United States. Clin Gastroenterol Hepatol. 2023;21(9):2418–2420.e3. doi:10.1016/j.cgh.2022.07.034

18. Shah J, Okubote T, Alkhouri N. Overview of Updated Practice Guidelines for Pediatric Nonalcoholic Fatty Liver Disease. Gastroenterol Hepatol (N Y). 2018;14(7):407–414.

19. Yu EL, Schwimmer JB. Epidemiology of Pediatric Nonalcoholic Fatty Liver Disease. Clin Liver Dis (Hoboken). 2021;17(3):196–199. doi:10.1002/cld.1027

20. Cowell E, Patel K, Heczey A, et al. Predisposing Conditions to Pediatric Hepatocellular Carcinoma and Association With Outcomes: Single-center Experience. J Pediatr Gastroenterol Nutr. 2019;68(5):695–699. doi:10.1097/MPG.0000000000002285

21. Zhang W, Sun B. Impact of age on the survival of patients with liver cancer: an analysis of 27,255 patients in the SEER database. Oncotarget. 2015;6(2):633–641.

22. Ren J, Tong YM, Cui RX, et al. Comparison of survival between adolescent and young adult vs older patients with hepatocellular carcinoma. World J Gastrointest Oncol. 2020;12(12):1394–1406. doi:10.4251/wjgo.v12.i12.1394

23. Hasegawa K, Kokudo N, Makuuchi M, et al. Comparison of resection and ablation for hepatocellular carcinoma: A cohort study based on a Japanese nationwide survey. Journal of Hepatology. 2013;58(4):724–729. doi:10.1016/j.jhep.2012.11.009

24. Lee YT, Wang JJ, Luu M, et al. The Mortality and Overall Survival Trends of Primary Liver Cancer in the United States. J Natl Cancer Inst. 2021;113(11):1531–1541. doi:10.1093/jnci/djab079

25. Kehm RD, Spector LG, Poynter JN, Vock DM, Altekruse SF, Osypuk TL. Does socioeconomic status account for racial and ethnic disparities in childhood cancer survival? Cancer. 2018;124(20):4090–4097. doi:10.1002/cncr.31560

26. Delavar A, Barnes JM, Wang X, Johnson KJ. Associations Between Race/Ethnicity and US Childhood and Adolescent Cancer Survival by Treatment Amenability. JAMA Pediatrics. 2020;174(5):428–436. doi:10.1001/jamapediatrics.2019.6074

27. Mathur AK, Osborne NH, Lynch RJ, Ghaferi AA, Dimick JB, Sonnenday CJ. Racial/Ethnic Disparities in Access to Care and Survival for Patients With Early-Stage Hepatocellular Carcinoma. Archives of Surgery. 2010;145(12):1158–1163. doi:10.1001/archsurg.2010.272

28. Kahla JA, Siegel DA, Dai S, et al. Incidence and 5-year survival of children and adolescents with hepatoblastoma in the United States. Pediatr Blood Cancer. 2022;69(10):e29763. doi:10.1002/pbc.29763

29. Wadhwani SI, Ge J, Gottlieb L, et al. Racial/Ethnic Disparities in Wait List Outcomes Are Only Partly Explained by Socioeconomic Deprivation Among Children Awaiting Liver Transplantation. Hepatology. 2022;75(1):115–124. doi:10.1002/hep.32106

30. Ebel NH, Lai JC, Bucuvalas JC, Wadhwani SI. A Review of Racial, Socioeconomic, and Geographic Disparities in Pediatric Liver Transplantation. Liver Transpl. 2022;28(9):1520–1528. doi:10.1002/lt.26437

31. Zhou K, Pickering TA, Gainey CS, et al. Presentation, Management, and Outcomes Across the Rural-Urban Continuum for Hepatocellular Carcinoma. JNCI Cancer Spectr. 2020;5(1):pkaa100. doi:10.1093/jncics/pkaa100

32. Glavas D, Bao QR, Scarpa M, et al. Treatment and Prognosis of Fibrolamellar Hepatocellular Carcinoma: a Systematic Review of the Recent Literature and Meta-analysis. J Gastrointest Surg. 2023;27(4):705–715. doi:10.1007/s11605-023-05621-z

33. Eggert T, McGlynn KA, Duffy A, Manns MP, Greten TF, Altekruse SF. Fibrolamellar hepatocellular carcinoma in the USA, 2000–2010: A detailed report on frequency, treatment and outcome based on the Surveillance, Epidemiology, and End Results database. United European Gastroenterol J. 2013;1(5):351–357. doi:10.1177/2050640613501507

34. Stipa F, Yoon SS, Liau KH, et al. Outcome of patients with fibrolamellar hepatocellular carcinoma. Cancer. 2006;106(6):1331–1338. doi:10.1002/cncr.21703

35. Weeda VB, Murawski M, McCabe AJ, et al. Fibrolamellar variant of hepatocellular carcinoma does not have a better survival than conventional hepatocellular carcinoma--results and treatment recommendations from the Childhood Liver Tumour Strategy Group (SIOPEL) experience. Eur J Cancer. 2013;49(12):2698–2704. doi:10.1016/j.ejca.2013.04.012

36. Bach PB, Pham HH, Schrag D, Tate RC, Hargraves JL. Primary Care Physicians Who Treat Blacks and Whites. New England Journal of Medicine. 2004;351(6):575–584. doi:10.1056/NEJMsa040609

