## Supplemental Material for "Incidence and survival of pediatric and adult hepatocellular carcinoma, United States, 2001-2020"

| **eTable 1. Incidence of hepatocellular carcinoma** | | |
| --- | --- | --- |
| **Total** |  |  |
| Age, years | **Count** | **Rate per 100,000 (95% CI)** |
| 0-19 | 813 | 0.056 (0.052-0.060) |
| 20+ | 354536 | 7.793 (7.767-7.819) |
| 0-14 | 383 | 0.036 (0.033-0.04) |
| 15-19 | 430 | 0.115 (0.104-0.126) |
| 20-29 | 1304 | 0.172 (0.163-0.182) |
| 30-39 | 3027 | 0.425 (0.410-0.440) |
| 40-64 | 181614 | 8.815 (8.774-8.856) |
| 65 + | 168591 | 22.084 (21.979-22.191) |
| **Stage** |  |  |
| **Localized** | |  |
| 0-19 | 299 | 0.021 (0.018-0.023) |
| 20+ | 168952 | 3.707 (3.690-3.725) |
| 0-14 | 149 | 0.014 (0.012-0.016) |
| 15-19 | 150 | 0.040 (0.034-0.047) |
| 20-29 | 529 | 0.070 (0.064-0.076) |
| 30-39 | 1353 | 0.190 (0.180-0.201) |
| 40-64 | 86162 | 4.173 (4.145-4.201) |
| 65 + | 80908 | 10.584 (10.510-10.657) |
| **Regional** | |  |
| 0-19 | 247 | 0.017 (0.015-0.019) |
| 20+ | 88873 | 1.948 (1.935-1.961) |
| 0-14 | 114 | 0.011 (0.009-0.013) |
| 15-19 | 133 | 0.036 (0.030-0.042) |
| 20-29 | 351 | 0.046 (0.042-0.052) |
| 30-39 | 783 | 0.110 (0.102-0.118) |
| 40-64 | 47378 | 2.300 (2.280-2.321) |
| 65 + | 40361 | 5.277 (5.226-5.330) |
| **Distant** |  |  |
| 0-19 | 234 | 0.016 (0.014-0.018) |
| 20+ | 51587 | 1.137 (1.127-1.147) |
| 0-14 | 102 | 0.010 (0.008-0.012) |
| 15-19 | 132 | 0.035 (0.030-0.042) |
| 20-29 | 312 | 0.041 (0.037-0.046) |
| 30-39 | 615 | 0.086 (0.079-0.093) |
| 40-64 | 26743 | 1.302 (1.286-1.317) |
| 65 + | 23917 | 3.141 (3.101-3.182) |
| **Histology** |  |  |
| **HCC (non-fHCC)** |  |  |
| 0-19 | 492 | 0.034 (0.031-0.037) |
| 20+ | 353648 | 7.772 (7.746-7.798) |
| 0-14 | 272 | 0.026 (0.023-0.029) |
| 15-19 | 220 | 0.059 (0.051-0.067) |
| 20-29 | 996 | 0.132 (0.124-0.140) |
| 30-39 | 2904 | 0.408 (0.394-0.423) |
| 40-64 | 181380 | 8.803 (8.762-8.844) |
| 65 + | 168368 | 22.055 (21.949-22.161) |
| **Fibrolamellar HCC** | |  |
| 0-19 | 321 | 0.022 (0.020-0.024) |
| 20+ | 888 | 0.021 (0.02-00.023) |
| 0-14 | 111 | 0.010 (0.009-0.013) |
| 15-19 | 210 | 0.056 (0.049-0.064) |
| 20-29 | 308 | 0.041 (0.036-0.046) |
| 30-39 | 123 | 0.017 (0.014-0.020) |
| 40-64 | 234 | 0.012 (0.011-0.014) |
| 65 + | 223 | 0.029 (0.026-0.034) |
| **Metropolitan status** | |  |
| **Metropolitan Counties** | |  |
| 0-19 | 710 | 0.057 (0.053-0.062) |
| 20+ | 305514 | 8.125 (8.096-8.154) |
| 0-14 | 330 | 0.040 (0.033-0.041) |
| 15-19 | 380 | 0.120 (0.108-0.132) |
| 20-29 | 1139 | 0.174 (0.164-0.185) |
| 30-39 | 2731 | 0.443 (0.426-0.460) |
| 40-64 | 157353 | 9.134 (9.089-9.180) |
| 65+ | 144291 | 23.167 (23.047-23.288) |
| **Nonmetropolitan Counties** | |  |
| 0-19 | 102 | 0.049 (0.040-0.060) |
| 20+ | 46904 | 6.203 (6.146-6.260) |
| 0-14 | 52 | 0.035 (0.026-0.046) |
| 15-19 | 50 | 0.090 (0.068-0.121) |
| 20-29 | 161 | 0.166 (0.141-0.193) |
| 30-39 | 279 | 0.307 (0.272-0.346) |
| 40-64 | 23334 | 7.210 (7.116-7.305) |
| 65+ | 23130 | 17.121 (16.90-17.345) |
| **Sex** |  |  |
| **Male** |  |  |
| 0-19 | 431 | 0.058 (0.053-0.064) |
| 20+ | 273079 | 12.891 (12.842-12.941) |
| 0-14 | 219 | 0.040 (0.035-0.046) |
| 15-19 | 212 | 0.111 (0.096-0.127) |
| 20-29 | 791 | 0.206 (0.192-0.220) |
| 30-39 | 2191 | 0.616 (0.590-0.642) |
| 40-64 | 149793 | 14.978 (14.902-15.055) |
| 65+ | 120304 | 35.768 (35.563-35.973) |
| **Female** |  |  |
| 0-19 | 382 | 0.054 (0.048-0.059) |
| 20+ | 81457 | 3.363 (3.339-3.386) |
| 0-14 | 164 | 0.032 (0.027-0.037) |
| 15-19 | 218 | 0.120 (0.104-0.137) |
| 20-29 | 513 | 0.138 (0.127-0.151) |
| 30-39 | 836 | 0.234 (0.218-0.250) |
| 40-64 | 31821 | 3.002 (2.968-3.035) |
| 65+ | 48287 | 11.332 (11.230-11.434) |
| **Race and Ethnicity** |  |  |
| **NH White** | |  |
| 0-19 | 444 | 0.056 (0.051-0.061) |
| 20+ | 205091 | 6.150 (6.123-6.177) |
| 0-14 | 200 | 0.035 (0.030-0.040) |
| 15-19 | 244 | 0.117 (0.103-0.133) |
| 20-29 | 594 | 0.141 (0.130-0.152) |
| 30-39 | 971 | 0.235 (0.221-0.250) |
| 40-64 | 99148 | 6.831 (6.788-6.874) |
| 65+ | 104378 | 17.843 (17.735-17.953) |
| **NH Black** | |  |
| 0-19 | 106 | 0.048 (0.039-0.058) |
| 20+ | 52670 | 10.819 (10.724-10.914) |
| 0-14 | 47 | 0.029 (0.022-0.039) |
| 15-19 | 59 | 0.103 (0.078-0.132) |
| 20-29 | 267 | 0.248 (0.219-0.279) |
| 30-39 | 775 | 0.830 (0.772-0.890) |
| 40-64 | 32768 | 13.878 (13.727-14.029) |
| 65+ | 18860 | 26.418 (26.036-26.805) |
| **NH American Indian/Alaska Native** | |  |
| 0-19 | ~ | ~ |
| 20+ | 3917 | 13.439 (13.002-13.886) |
| 0-14 | ~ | ~ |
| 15-19 | ~ | ~ |
| 20-29 | 8 | 0.116 (0.050-0.228) |
| 30-39 | 28 | 0.497 (0.330-0.719) |
| 40-64 | 2322 | 15.383 (14.760-16.026) |
| 65+ | 1559 | 38.116 (36.185-40.127) |
| **NH Asian or Pacific Islander** | |  |
| 0-19 | 52 | 0.069 (0.051-0.090) |
| 20+ | 28918 | 14.706 (14.533-14.881) |
| 0-14 | 29 | 0.052 (0.035-0.074) |
| 15-19 | 23 | 0.120 (0.076-0.179) |
| 20-29 | 156 | 0.314 (0.267-0.368) |
| 30-39 | 748 | 1.512 (1.405-1.625) |
| 40-64 | 13304 | 13.357 (13.130-13.587) |
| 65+ | 14710 | 48.718 (47.923-49.523) |
| **Hispanic** | |  |
| 0-19 | 185 | 0.058 (0.050-0.067) |
| 20+ | 56815 | 14.042 (13.922-14.162) |
| 0-14 | 97 | 0.040 (0.032-0.048) |
| 15-19 | 88 | 0.113 (0.091-0.140) |
| 20-29 | 244 | 0.159 (0.140-0.180) |
| 30-39 | 434 | 0.317 (0.287-0.348) |
| 40-64 | 30612 | 13.915 (13.759-14.072) |
| 65+ | 25525 | 45.224 (44.662-45.791) |
| **Socioeconomic Status** | |  |
| **Top 25% Economic Status** | |  |
| 0-19 | 295 | 0.063 (0.056-0.071) |
| 20+ | 106,570 | 7.296 (7.252-7.341) |
| 0-14 | 140 | 0.041 (0.034-0.048) |
| 15-19 | 155 | 0.130 (0.110-0.152) |
| 20-29 | 422 | 0.178 (0.161-0.196) |
| 30-39 | 1,044 | 0.437 (0.411-0.464) |
| 40-64 | 52,432 | 7.666 (7.600-7.733) |
| 65+ | 52,672 | 22.025 (21.836-22.215) |
| **Middle 25-75% Economic Status** | |  |
| 0-19 | 415 | 0.053 (0.048-0.058) |
| 20+ | 201,762 | 8.139 (8.104-8.175) |
| 0-14 | 188 | 0.033 (0.028-0.038) |
| 15-19 | 227 | 0.111 (0.097-0.127) |
| 20-29 | 742 | 0.176 (0.164-0.189) |
| 30-39 | 1,593 | 0.414 (0.394-0.435) |
| 40-64 | 105,433 | 9.508 (9.45-9.567) |
| 65+ | 93,994 | 22.382 (22.238-22.526) |
| **Bottom 25% Economic Status** | |  |
| 0-19 | 71 | 0.050 (0.039-0.064) |
| 20+ | 35,718 | 8.310 (8.223-8.397) |
| 0-14 | 40 | 0.039 (0.028-0.053) |
| 15-19 | 31 | 0.085 (0.058-0.121) |
| 20-29 | 104 | 0.153 (0.125-0.185) |
| 30-39 | 283 | 0.467 (0.414-0.525) |
| 40-64 | 18,445 | 9.858 (9.713-10.003) |
| 65+ | 16,886 | 22.463 (22.123-22.806) |

Rates per 100,000 persons. Three decimal places were used for pediatric ages instead of expressing rates per 1 million. Abbreviations: fibrolamellar hepatocellular carcinoma (fHCC), hepatocarcinoma (HCC), Non-Hispanic (NH), confidence intervals (CI), reference group (ref), not applicable (~). The left column indicates year of age.

| **eTable 2. Relative risk of hepatocellular carcinoma incidence** | | | |  |
| --- | --- | --- | --- | --- |
|  | **0-19 years model** | | **20+ years model** | |
| **Variable** | **RR (95%CI)** | **p-value** | **RR (95%CI)** | **p-value** |
| **Metro 250K-1M vs Metro >1M** | 0.97 (0.81-1.17) | 0.7662 | 0.95 (0.89-1.01) | 0.0917 |
| **Metro <250K vs Metro >1M** | 0.76 (0.56-1.03) | 0.074 | 0.88 (0.82-0.95) | 0.0004 |
| **Non-Metro vs Metro >1M** | 0.88 (0.69-1.13) | 0.3151 | 0.83 (0.78-0.89) | <.0001 |
| **Bottom 25% vs Top 25%** | 0.89 (0.66-1.18) | 0.4123 | 1.09 (1.02-1.17) | 0.0076 |
| **25-75% vs Top 25%** | 0.88 (0.77-1.03) | 0.1098 | 1.05 (0.99-1.11) | 0.1055 |
| **15-19 years vs 0-14 years** | 3.15 (2.73-3.64) | <.0001 | ~ | ~ |
| **30-39 years vs 20-29 years** | ~ | ~ | 2.42 (2.18-2.69) | <.0001 |
| **40-64 years vs 20-29 years** | ~ | ~ | 55.27 (50.36-60.65) | <.0001 |
| **years vs 20-29 years** | ~ | ~ | 154.56 (140.79-169.68) | <.0001 |

Percentages indicate economic status (e.g. Bottom 25%). Abbreviations: Relative Risk (RR), metropolitan (metro).

| **eTable 3. 5-year relative survival of patients with hepatocellular carcinoma** | |  |
| --- | --- | --- |
| **Total** |  |  |
| Age, years | **Count** | **5 Year RS % (95% CI)** |
| 0-19 | 702 | 46.4 (42.4-50.3) |
| 20+ | 256,704 | 20.7 (20.5-20.2) |
| 0-14 | 334 | 48.5 (42.8-54.0) |
| 15-19 | 368 | 43.6 (38.1-49.0) |
| 20-29 | 1129 | 44.4 (41.4-47.0) |
| 30-39 | 2,581 | 36.1 (34.2-38.3) |
| 40-64 | 142,595 | 22.7 (22.5-23.2) |
| 65 + | 110,399 | 17.0 (16.7-17.1) |
| **Total 2001-2007** | |  |
| 0-19 | 256 | 37.6 (31.7-43.5) |
| 20+ | 62,722 | 16.4 (16.1-16.7) |
| 0-14 | 118 | 38.2 (29.5-46.9) |
| 15-19 | 138 | 37.1 (29.1-45.1) |
| 20-29 | 372 | 33.8 (29.0-38.6) |
| 30-39 | 960 | 30.8 (27.9-33.7) |
| 40-64 | 34,578 | 19.9 (19.0-19.9) |
| 65 + | 26,812 | 11.5 (11.0-11.9) |
| **Total 2008-2020** | |  |
| 0-19 | 446 | 52.3 (30.1-57.4) |
| 20+ | 193,982 | 22.2 (22.0-22.4) |
| 0-14 | 216 | 58.4 (50.9-65.1) |
| 15-19 | 230 | 46.3 (38.9-53.5) |
| 20-29 | 757 | 50.6 (46.6-54.4) |
| 30-39 | 1,621 | 39.4 (36.7-42.0) |
| 40-64 | 108,017 | 23.8 (23.5-24.1) |
| 65 + | 83,587 | 19.1 (18.7-19.5) |
| **Stage** |  |  |
| **Localized** | |  |
| 0-19 | 255 | 75.1 (68.8-80.4) |
| 20+ | 121,569 | 33.6 (33.3-14.9) |
| 0-14 | 128 | 79.0 (70.1-19.6) |
| 15-19 | 127 | 71.4 (61.7-15.0) |
| 20-29 | 455 | 71.4 (66.6-16.1) |
| 30-39 | 1107 | 57.8 (54.6-14.4) |
| 40-64 | 67,560 | 36.9 (36.5-37.3) |
| 65 + | 52,447 | 27.7 (27.2-18.3) |
| **Regional** | |  |
| 0-19 | 204 | 44.5 (37.1-51.6) |
| 20+ | 65,784 | 12.0 (11.7-5.2) |
| 0-14 | 90 | 49.6 (38.2-14.5) |
| 15-19 | 114 | 40.4 (30.7-5.1) |
| 20-29 | 298 | 34.8 (29.0-10.8) |
| 30-39 | 688 | 23.0 (19.7-4.9) |
| 40-64 | 37,815 | 13.0 (12.6-13.4) |
| 65 + | 26,983 | 9.70 (9.3-13.1) |
| **Distant** |  |  |
| 0-19 | 215 | 14.1 (9.5-28.2) |
| 20+ | 39,568 | 3.5 (3.2-33.6) |
| 0-14 | 101 | 14.0 (7.6-22.7) |
| 15-19 | 114 | 15.0 (8.9-23.0) |
| 20-29 | 288 | 15.0 (11.0-8.2) |
| 30-39 | 544 | 11.5 (8.9-31.1) |
| 40-64 | 22,062 | 3.3 (3.1-34.0) |
| 65 + | 16,674 | 3.1 (2.8-8.0) |
| **Histology** |  |  |
| **HCC (non-fHCC)** |  |  |
| 0-19 | 440 | 44.8 (39.8-49.7) |
| 20+ | 255,990 | 20.7 (20.5-20.9) |
| 0-14 | 245 | 49.5 (42.8-55.9) |
| 15-19 | 195 | 38.5 (31.1-45.9) |
| 20-29 | 861 | 47.0 (43.4-50.5) |
| 30-39 | 2,475 | 35.7 (33.7-37.8) |
| 40-64 | 142,411 | 22.7 (22.5-23.0) |
| 65 + | 110,243 | 17.0 (16.7-17.3) |
| **Fibrolamellar HCC** | |  |
| 0-19 | 262 | 49.3 (42.7-55.6) |
| 20+ | 714 | 35.3 (31.9-39.8) |
| 0-14 | 89 | 52.5 (40.8-62.9) |
| 15-19 | 173 | 47.8 (39.6-55.5) |
| 20-29 | 268 | 36.0 (30.2-42.6) |
| 30-39 | 106 | 44.8 (34.2-54.8) |
| 40-64 | 184 | 37.7 (30.1-45.3) |
| 65 + | 156 | 26.6 (18.3-35.7) |
| **Metropolitan Status** | |  |
| **Metropolitan Counties** | |  |
| 0-19 | 611 | 47.3 (43.0-51.5) |
| 20+ | 223,838 | 21.3 (21.1-21.5) |
| 0-14 | 287 | 51.4 (45.1-57.3) |
| 15-19 | 324 | 43.8 (37.9-49.5) |
| 20-29 | 999 | 45.3 (41.9-48.6) |
| 30-39 | 2,348 | 35.7 (33.6-37.8) |
| 40-64 | 124,715 | 23.3 (23.0-23.5) |
| 65+ | 95,776 | 17.6 (17.2-17.9) |
| **Nonmetropolitan Counties** | |  |
| 0-19 | 91 | 40.7 (30.1-51.0) |
| 20+ | 32,814 | 16.9 (16.4-17.4) |
| 0-14 | 47 | 43.6 (28.6-57.6) |
| 15-19 | 44 | 37.4 (22.8-52.0) |
| 20-29 | 130 | 36.5 (27.7-45.4) |
| 30-39 | 232 | 40.2 (33.5-46.9) |
| 40-64 | 17,847 | 18.8 (18.2-19.5) |
| 65+ | 14,605 | 13.5 (12.7-14.3) |
| **Sex** |  |  |
| **Male** |  |  |
| 0-19 | 381 | 46.8 (41.4-52.0) |
| 20+ | 198,821 | 19.9 (19.7-20.1) |
| 0-14 | 197 | 51.8 (44.2-58.9) |
| 15-19 | 184 | 41.6 (34.0-49.0) |
| 20-29 | 695 | 39.6 (35.7-43.5) |
| 30-39 | 1,887 | 33.1 (30.8-35.3) |
| 40-64 | 118,494 | 21.4 (21.1-21.7) |
| 65+ | 77,745 | 16.5 (16.2-16.8) |
| **Female** |  |  |
| 0-19 | 321 | 46.0 (40.0-51.7) |
| 20+ | 57,883 | 23.7 (23.3-24.1) |
| 0-14 | 137 | 48.1 (38.9-56.6) |
| 15-19 | 184 | 44.3 (36.3-51.9) |
| 20-29 | 434 | 52.0 (46.7-56.9) |
| 30-39 | 694 | 44.5 (40.4-48.4) |
| 40-64 | 24,101 | 29.4 (28.7-30.0) |
| 65+ | 32,654 | 18.2 (17.7-18.8) |
| **Race and Ethnicity** |  |  |
| **NH White** | |  |
| 0-19 | 372 | 48.9 (43.3-54.1) |
| 20+ | 142392 | 19.8 (19.5-20.0) |
| 0-14 | 170 | 53.2 (45.0-60.7) |
| 15-19 | 202 | 45.2 (37.8-52.4) |
| 20-29 | 491 | 48.2 (43.3-53.0) |
| 30-39 | 822 | 42.9 (39.2-46.6) |
| 40-64 | 75,998 | 22.5 (22.2-22.9) |
| 65+ | 65081 | 15.4 (15.1-15.8) |
| **NH Black** | |  |
| 0-19 | 93 | 41.1 (30.4-51.5) |
| 20+ | 39,152 | 17.3 (16.9-17.8) |
| 0-14 | 43 | 49.6 (33.0-64.3) |
| 15-19 | 50 | 34.1 (20.8-48.0) |
| 20-29 | 241 | 30.7 (24.8-36.9) |
| 30-39 | 670 | 24.2 (20.8-27.7) |
| 40-64 | 25,815 | 17.1 (16.6-17.6) |
| 65+ | 12,426 | 16.9 (16.0-17.8) |
| **NH Indian or Alaska Native** |  |  |
| 0-19 | ~ | ~ |
| 20+ | 2,550 | 17.0 (15.3-18.9) |
| 0-14 | ~ | ~ |
| 15-19 | ~ | ~ |
| 20-29 | 7 | 18.1 (0.8%-54.3) |
| 30-39 | 17 | 40.2 (17.2-62.4) |
| 40-64 | 1,594 | 18.7 (16.5-21.1) |
| 65+ | 932 | 13.4 (10.5-16.5) |
| **NH Asian or Pacific Islander** |  |  |
| 0-19 | 44 | 41.0 (25.7-55.7) |
| 20+ | 21971 | 31.4 (30.7-32.1) |
| 0-14 | 25 | 43.3 (22.8-62.4) |
| 15-19 | 19 | 38.0 (16.4-59.6) |
| 20-29 | 140 | 36.3 (28.1-44.7) |
| 30-39 | 637 | 34.1 (30.3-38.0) |
| 40-64 | 10,712 | 34.9 (33.9-35.9) |
| 65+ | 10,482 | 27.1 (26.1-28.2) |
| **Hispanic** | |  |
| 0-19 | 170 | 43.1 (34.9-51.0) |
| 20+ | 44,990 | 20.8 (20.4-21.3) |
| 0-14 | 88 | 46.8 (35.3-57.4) |
| 15-19 | 82 | 38.4 (26.7-50.0) |
| 20-29 | 219 | 56.7 (49.3-63.4) |
| 30-39 | 377 | 45.0 (39.6-50.3) |
| 40-64 | 25,496 | 23.2 (22.6-23.8) |
| 65+ | 18,898 | 16.1 (15.5-16.8) |
| **Socioeconomic Status** |  |  |
| **Top 25% Economic Status** | |  |
| 0-19 | 414 | 44.6 (39.4-49.6) |
| 20+ | 168,872 | 19.7 (19.5-19.9) |
| 0-14 | 191 | 51.9 (44.2-59.1) |
| 15-19 | 223 | 38.1 (31.2-45.0) |
| 20-29 | 729 | 45.3 (41.4-49.2) |
| 30-39 | 1,594 | 34.8 (32.2-37.3) |
| 40-64 | 95,327 | 21.5 (21.2-21.8) |
| 65+ | 71,222 | 16.2 (15.9-16.6) |
| **Middle 25-75% Economic Status** | |  |
| 0-19 | 102 | 54.1 (43.5-63.6) |
| 20+ | 29,723 | 25.6 (25.0-26.1) |
| 0-14 | 53 | 47.2 (32.6-60.5) |
| 15-19 | 49 | 61.6 (45.9-73.9) |
| 20-29 | 119 | 42.2 (32.5-51.6) |
| 30-39 | 336 | 40.8 (35.2-46.4) |
| 40-64 | 15,382 | 29.1 (28.3-29.9) |
| 65+ | 13,886 | 20.4 (19.6-21.3) |
| **Bottom 25% Economic Status** | |  |
| 0-19 | 155 | 46.7 (38.1-54.9) |
| 20+ | 51,635 | 20.9 (20.5-21.3) |
| 0-14 | 76 | 46.5 (34.2-57.9) |
| 15-19 | 79 | 47.0 (34.8-58.2) |
| 20-29 | 252 | 43.4 (36.9-49.7) |
| 30-39 | 567 | 36.0 (31.9-40.1) |
| 40-64 | 28,218 | 23.0 (22.4-23.5) |
| 65+ | 22,598 | 17.2 (16.5-17.8) |

Abbreviations: Relative Survival (RS) confidence intervals (CI) 0-19 years of age (0-19), 20 years or older (20+), fibrolamellar hepatocarcinoma (fHCC), hepatocarcinoma (HCC), Non-Hispanic (NH). The left column indicates years of age.

**eFigure 1** **5-year overall survival of adult (left) and pediatric (right) patients with hepatocellular carcinoma.** A. Diagnosis year. B. Stage. C. Histology. Abbreviation: fibrolamellar hepatocellular carcinoma (fHCC), hepatocarcinoma (HCC).

**eFigure 2** **5-year overall survival of adult (left) and pediatric (right) patients with hepatocellular carcinoma**. A. Sex. B. Race and ethnicity. C. Socioeconomic status by county. D. Metropolitan status. Abbreviation: metropolitan (metro), Non-Hispanic (NH).
